# The utility of 8-Hydroxydeoxyguanosine and 8-Oxoguanine DNA glycosylase as a novel diagnostic marker for autistic children

**DOI:** 10.1101/2025.01.15.25320384

**Authors:** Hawnaz Mohammad Ismael, Parween Abdulsamad Ismail

## Abstract

Autism spectrum disorder (ASD) is a neurodevelopmental condition that develops in childhood. Although genetics are a key factor in its development, neurological, environmental, and immunological influences also play significant roles. This study investigated the oxidative DNA damage (ODD) biomarker 8-hydroxy-2-deoxyguanosine (8OHdG) in children with autism, exploring its connection to disease severity and the DNA repair enzyme oxoguanine glycosylase 1 (OGG1). This study investigated the oxidative DNA damage (ODD) biomarker, 8-hydroxy-2-deoxyguanosine (8OHdG), in children with autism, examining its relationship with disease severity and the DNA repair enzyme oxoguanine glycosylase 1 (OGG1). The study included 89 children with ASD and 29 typically developing children in an observational controlled cross-sectional design. Autism severity was assessed using the Diagnostic and Statistical Manual of Mental Disorders (DSM-5). ELISA was used to measure serum levels of 8-OHdG and OGG1.The results showed that children with autism had significantly higher serum 8-OHdG levels compared to healthy children (P=0.04), with a significant positive correlation with autism severity (P=0.02). Additionally, serum OGG1 levels were significantly lower in children with autism than in their healthy counterparts (P=0.0004), with a notable positive association with disease severity (P=0.0001). These findings indicate that elevated serum 8-OHdG levels may play a key role in oxidative DNA damage in ASD. Higher levels of 8-OHdG in children with severe ASD symptoms could serve as a potential biomarker for diagnosis. Furthermore, the reduced levels of the repair enzyme OGG1, associated with increased 8-OHdG levels, contribute to the observed DNA damage in ASD.

## Introduction

Autism Spectrum Disorder (ASD) is a neurodevelopmental condition impacting social, verbal, and nonverbal communication, characterized by repetitive behaviors and restricted interests(1). The global prevalence of ASD in children is over 1 in 100 with a male-to-female ratio of 4:2(2). Childhood autism is the most acute form of ASD. It is generally diagnosed between the ages of 6 and 10 and is more commonly found in males(3). The etiology of autism are linked to genetic, neurological, and environmental influences. Key pathways that connect these risk factors include oxidative stress and neurotransmitters. (4).

DNA damage can result from an imbalance between oxidants and antioxidants, playing a role in ASD pathogenesis(5). Reactive Oxygen Species (ROS) can cause various DNA damages, particularly targeting the base Guanine (G) due to its low redox potential. This leads to several oxidized G products in DNA and the ribonucleotide pool(6).

In both nuclear and mitochondrial DNA, 8-hydroxy-2-deoxyguanosine (8-OHdG) is a prevalent form of oxidative damage caused by free radicals, known for its mutagenic properties(7). It has been established as a significant oxidative stress biomarker, with levels measured in human organs, leukocyte DNA, and urine(8). For example, a recent study found that autistic individuals have notably higher urinary 8-OHdG levels compared to non-autistic individuals(5), suggesting a possible link to cerebellar impairment in autism spectrum disorder (ASD)(9).

The base excision repair (BER) pathway is responsible for fixing 8-oxoG and other base modifications(10). Studies indicate that 8-oxoguanine DNA glycosylase (OGG1) is multifaceted in its functions beyond merely repairing DNA damage. Besides its glycosylase role in fixing 8-oxoG lesions, OGG1 also plays a part in regulating epigenetic modifications. It accomplishes this by attracting epigenetic modifiers, which then either activate or suppress gene transcription, including those genes influenced by the estrogen receptor(11, 12). OGG1 aids in repairing DNA damage caused by reactive oxygen species (ROS), which are associated with various brain function abnormalities, including autism(13). Research indicates that both autistic patients and autism mouse models exhibit elevated levels of DNA damage and reduced efficiency in DNA repair mechanisms.(14).

The current study analyzes autistic children to neurotypical children in order to examine the association between the enzyme repair capacity of OGG1 and the serum levels of the oxidative DNA damage biomarker 8-OHdG. This study aims to show how oxidative stress could be used as a diagnostic tool in the genesis of autism.

## Materials and methods

### Subjects

The case-control study involved 60 children with ASD (52 males and 8 females), aged between three and twenty years, from autism centers in Erbil city, Kurdistan Region - Iraq. The study was conducted from September 22, 2024, to November 4, 2024. For comparison, a control group of 29 healthy children (23 males and 6 females) was included. All ASD participants had a confirmed clinical diagnosis of ASD by qualified professionals based on the Diagnostic and Statistical Manual of Mental Disorders (DSM-IV). The children with ASD were further categorized into three subgroups: Level I, Level II, or Level III, with Level III requiring the most support.

The study adhered to the ethical standards of the responsible human experimentation committee and the latest version of the Declaration of Helsinki. Ethical approval was obtained from the Council of Ministry / Ministry of Higher Education & Scientific Research – Salahaddin University Directorate Postgraduate (approval no. 13793, issued on September 11, 2024), for all autism centers in Erbil city. Written informed consent was secured from the parents or legal guardians of the participating children, and the consent forms have been archived by the author.

### Method

A total of 3 ml of venous blood was collected from children with ASD and control groups using EDTA tubes. After coagulation, the samples were centrifuged for 20 minutes at 3000 rpm, and the serum was kept for later analysis at −20°C. ELISA was used to test plasma samples. Following the manufacturer’s instructions, ELISA kits were used to assess the levels of 8-hydroxy-2’-deoxyguanosine (8-OHdG) using an antigen-antibody sandwich approach. A microplate that had been pre-coated with particular antibodies was utilized for evaluation in a quantitative sandwich enzyme immunoassay. After the addition of the immobilized antibody bound to 8-OHdG, an enzyme-linked secondary antibody specific to 8-OHdG was added. At 450 nm, absorbance was measured. For OGG1, the same process was applied.

### Statistical Analysis

Statistical analyses were performed using GraphPad Prism 9.5.1 and MedCalc statistical software version 20.018. The Shapiro–Wilk normality test determined if the data followed a normal distribution. As the data were normally distributed, a Mann-Whitney test was used to compare differences between the control (CON) and ASD groups for each measurement (8-OHdG and OGG1). Additionally, one-way ANOVA assessed statistical differences between various stages of ASD and control groups for each measurement (8-OHdG and OGG1), with significance set at p < 0.05. All data are presented as Mean ± SEM.

The Receiver Operating Characteristic (ROC) curve was employed to assess the diagnostic accuracy of 8-OHdG and OGG1 for ASD. This curve was plotted to determine the cut-off point, considering p < 0.05 as statistically significant.

## Results

### Serum levels of 8-OHdG

The serum level of 8-hydroxydeoxyguanosine in ASD was found to be significantly increase (P=0.043) in Figure1 (448.2±28.3pg/ml) than in healthy controls (368.4±12.2pg/ml) with respect to the sex, age, ethnicity, and clinical characteristics.

**Figure 1.**
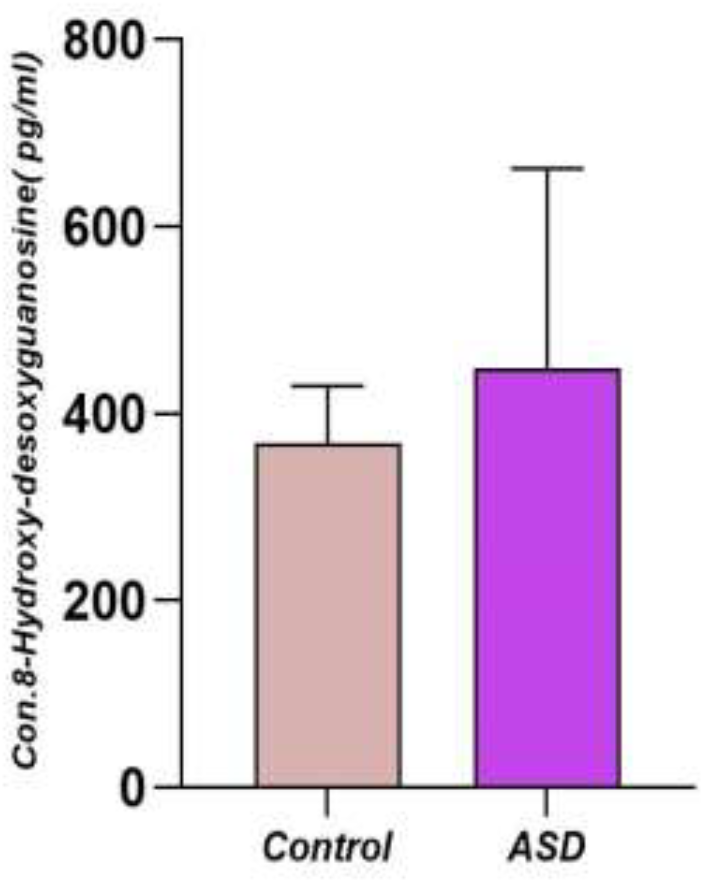
Comparison of 8-OHdG level in serum between ASD disease and control cases.

### Serum levels of 8-OHdG among ASD severity

With increasing autism severity, serum 8-OHdG levels rise significantly (p value = 0.029)in autistic individuals (Level I = 400.7±108.4 pg/ml),(Level II = 424.4 ±163.9 pg/ml) and (Level III= 539.1±324.9 pg/ml)compared to case control(373.7 ± 65.5) (Figure 2).

**Figure 2.**
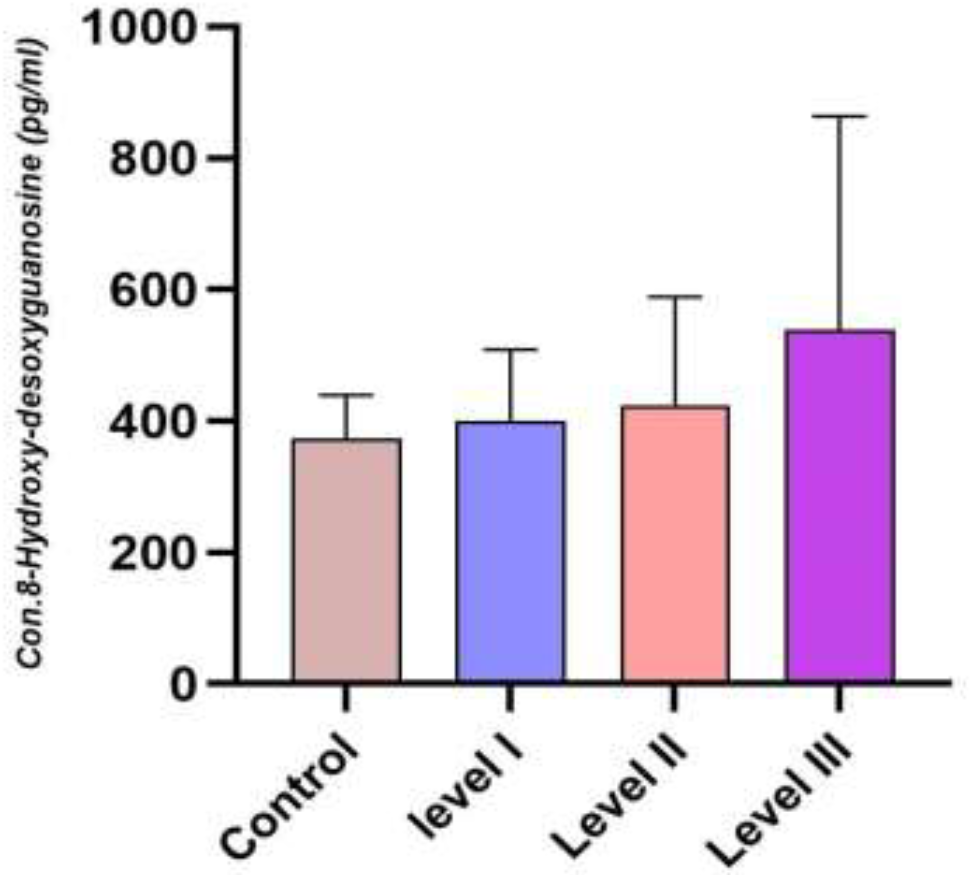
Levels of 8-OHdG according to disorder severity and control

### Serum levels of OGG1

As a key DNA repair enzyme, Serum OGG1mean level was lower than healthy group,with statistically highly significant differences(p = 0.0004). The mean level of the study group was (271.9 ± 23.4 pg/ml) whereas the control group means level was (371.9±13.6 pg/ml), as shown Figure 3.

**Figure 3.**
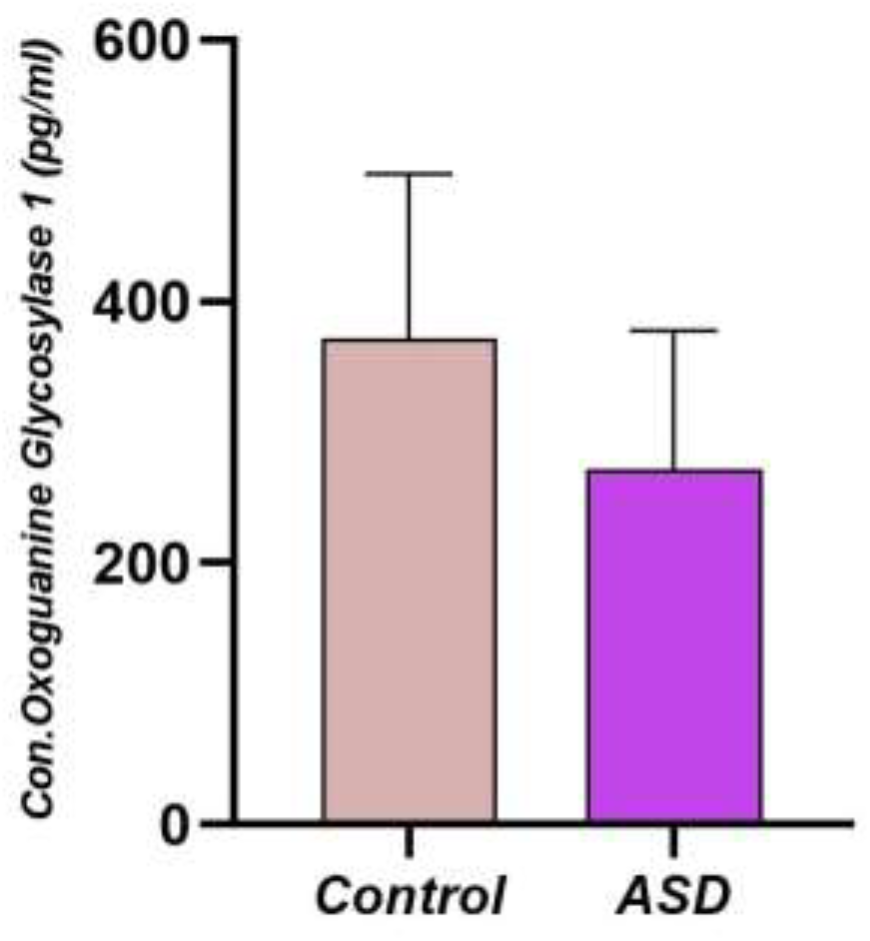
Comparison of OGG1 level in serum between ASD disease and control cases.

**Figure 4.**
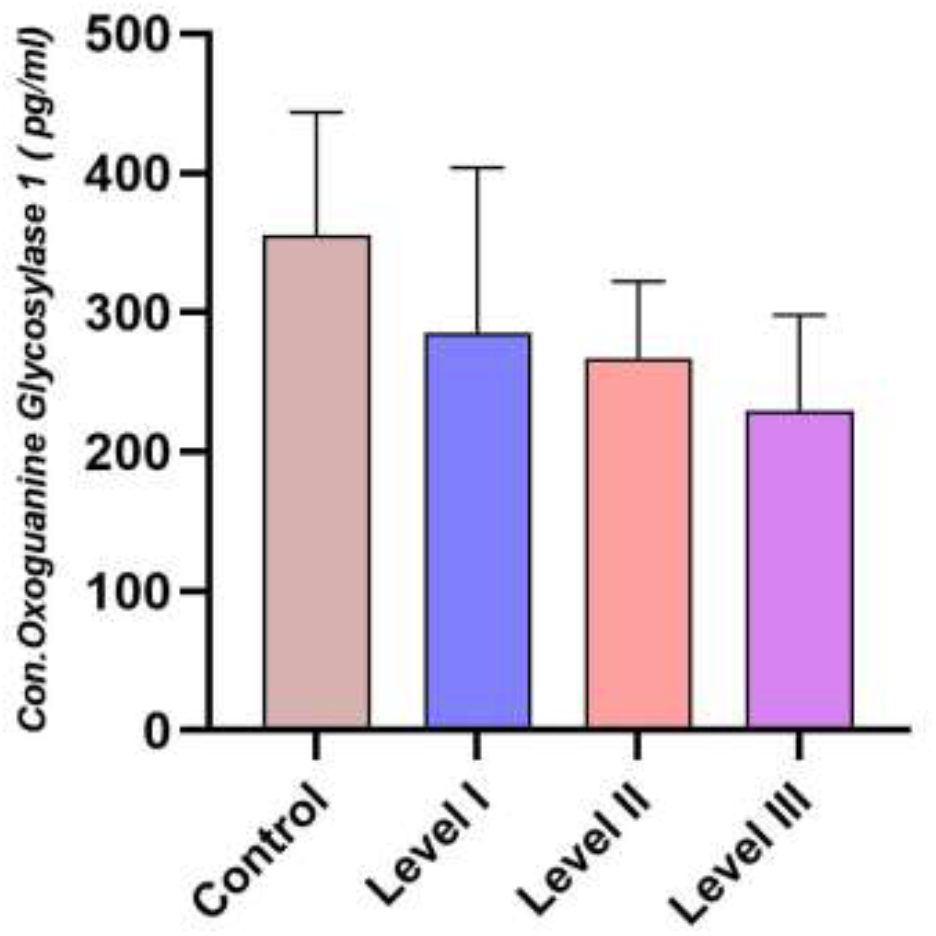
Levels of OGG1 according to disorder severity and control.

### Serum levels of OGG1 among ASD severity

Among the increasing autism severity, serum OGG1 levels rise significantly (p value = <0.0001)in autistic individuals (Level I = 285.5±118.6 pg/ml),(Level II = 267.4 ±55 pg/ml) and (Level III= 229.8±68.32),meanwhile mean serum of control groups (355.0± 88.69 pg/ml). Multiple linear regression analysis showed that there was significant association between the presence of OGG1 and the severity of childhood ASD compared to case control.

### ROC curve analysis

ROC analysis was conducted to evaluate the effectiveness of various factors that influenced dependent variables in diagnosing ASD. Table 5 demonstrates the AUCs, specificity, 95% CI, p-values, cut off values and sensitivity of all ASD patients compared to control subjects for each parameter (8OHdG,OGG1).The ROC curve analysis with AUC, sensitivity, and specificity estimations is summarized in Figure 5.

**Figure 5.**
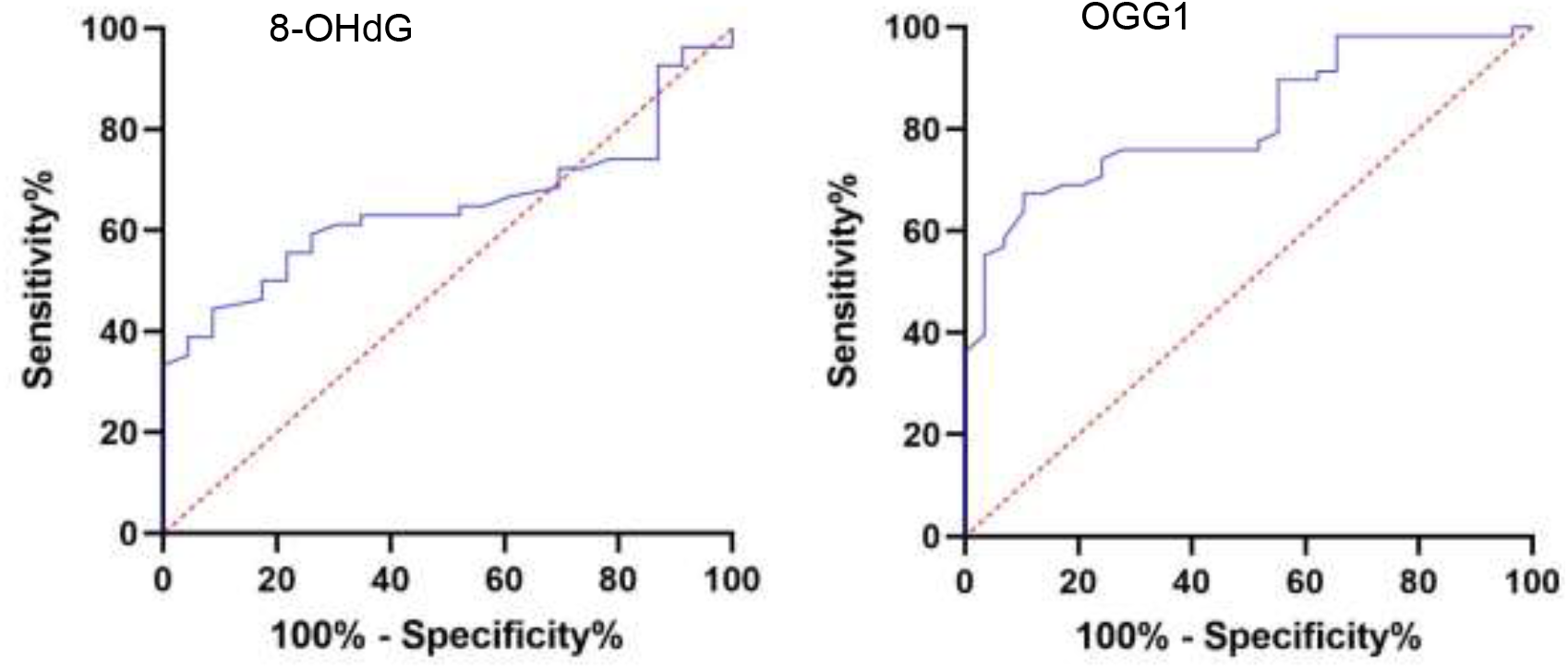
Receiver Operator Characteristics (ROC) curve evaluating the utility of 8-OHdG and OGG1 in the diagnosis of ASD.

**Table 1.** Receiver Operator Characteristics (ROC) curve evaluating the utility of 8-OHdG and OGG1 in the diagnosis of ASD.

| Parameter | AUC | 95% CI | P-value | Cut off Values | Sensitivity | Specificity |
| --- | --- | --- | --- | --- | --- | --- |
| 8-OHdG | 0.646 | (0.5262,0.7669) | 0.042 | 320.5 | 67.8 | 10.71 |
| OGG1 | 0.817 | (0.7136,0.8937) | 0.0001 | 276.1 | 67.2 | 89.6 |

## Discussion

Despite extensive research, the precise cause of autism spectrum disorder (ASD) remains unclear. Various factors, including genetic, epigenetic, and environmental elements, may influence its etiopathogenesis(15). Oxidative stress, known for its genotoxic properties, can cause both single- and double-stranded DNA breaks. The development of severe diseases, including ASD, is related to the extent of DNA damage and the efficiency of repair mechanisms(16). In vivo, ROS interacts with biopolymers such proteins, lipids, enzymes, and DNA, causing gene mutation, lipid peroxidation, protein denaturation, and enzyme inactivation(17). Oxidative stress signifies the detrimental impact of oxidative reactions in living organisms. Elevated oxidative stress levels are believed to exacerbate molecular biological oxidative damage, potentially contributing to various diseases and premature aging(18).

Oxidative DNA damage, a consequence of oxidative stress, often targets guanine nucleobases because they are more susceptible to oxidation(19). Excess ROS will hydroxylate guanine bases to generate 8-hydroxyguanine (8-OH-Gua), which will then either produce 8-hydroxy-2’-deoxyguanosine (8-OHdG) through electron abstraction or 8-oxo-7,8-dihydro-2’-deoxy-guanosine (8-oxodG) through keto-enol tautomerism of 8-OHdG. Nonetheless, in the majority of the literature, 8-OHdG and 8-oxodG are typically used interchangeably. Oxidative damage, one of the primary types of DNA damage that results in DNA alteration with changed functions, is caused by this production(20).These mutagenic deoxyguanosines are more frequently used as an indicator of oxidative damage and are further categorized as a biomarker in the early detection of various diseases because of their ease of cell membrane contacting(21).

Higher concentrations of 8OHdG have been found in the lymphocytes and cerebrospinal fluid of people with neurodegenerative disorders, such as Alzheimer’s and Parkinson’s diseases, and also in children who have suffered brain damage(22). This indicates that oxidative DNA damage (ODD) significantly contributes to the development of various neurological diseases. Recognizing ODD’s role in autism’s pathophysiology has spurred interest in identifying early biological markers for clinical, therapeutic, or preventive interventions.

n our research, we compared serum levels of the DNA hydroxylation biomarker 8-hydroxy-2-deoxyguanosine (8-OHdG) in children with autism to those in age- and sex-matched typically developing controls. We found that children with ASD had significantly higher serum 8-OHdG levels than the control group, effectively distinguishing between typically developing and autistic children. These results are consistent with prior studies that reported elevated 8-OHdG levels in the blood, urine, and post-mortem cerebellar tissue of autistic children(9). The increased 8-OHdG levels observed in autistic children may indicate abnormal reactive oxygen species (ROS) expression, which could affect neuronal differentiation, migration, and synaptic signaling.(23), and indicating higher susceptibility to oxidative stress and impaired DNA repair mechanisms.

The mechanisms that underlie neurodevelopmental disorders such as Autism Spectrum Disorders (ASD) may involve DNA damage initiated by reactive oxygen species (ROS), measured as 8-oxoguanine (8-oxoG), and epigenetic modifications of histone proteins and DNA that regulate gene expression. 8-oxoG is repaired by the enzyme oxoguanine glycosylase 1 (OGG1). Recently, both 8-oxoG and OGG1 have been identified as epigenetic markers that contribute to the regulation of gene expression, either increasing or decreasing it. The role of OGG1 as an enzyme that repairs DNA damage and an epigenetic regulator in reducing neurodevelopmental problems brought on by physiological and ethanol-induced increases in reactive oxygen species(24).

OGG1 is essential for normal brain development due to its role in DNA repair and as an epigenetic regulator. OGG1 deficiency has been associated with neurodevelopmental disorders(14). The study found that levels of OGG1 were notably lower in autistic children compared to non-autistic individuals, as depicted in Figure 3. This indicates that autistic children experience spontaneous DNA damage, evidenced by their lower baseline levels of OGG1 at all sampling points compared to healthy controls. These findings are consistent with Porokhovnik et al.’s research, which also observed baseline DNA damage in peripheral blood samples from mothers and infants with autism using the comet test(25).

In this study, patients with level III autism had significantly higher serum 8-OHdG levels than children with level I to level II autism. This suggests that the elevation of serum 8-OHdG levels is closely linked to the severity of autism. The increased neurological damage associated with higher autism severity may lead to greater production of 8-OHdG protein. However, it is challenging to determine whether the rise in serum 8-OHdG levels is merely a consequence of autism or plays a pathogenic role in the disease.

Studies reveal that oxidative stress is linked to DNA damage in autistic individuals. Additionally, there is evidence linking borderline personality disorder (BPD) individuals to elevated oxidative stress and DNA damage(26). Autistic patients’ post-mortem examinations have revealed substantial elevations in 8-oxo-2’-deoxyguanosine (8-oxo-dG), the primary DNA oxidation product.

(27). According to another study, methionine transfer from S-adenosylmethionine to DNA, RNA, proteins, phospholipids, and neurotransmitters may be hampered in autistic children due to decreased methylation ability and increased oxidative stress(28).

Owing to research, mast cells are activated by oxidative stress, which causes them to produce proinflammatory and neurosensitizing chemicals such bradykinin, NO, IL-6, and histamine. These chemicals have the potential to damage the blood-gut and blood-brain barriers.(29). Based on certain research, enterotoxic chemicals may infiltrate autistic individuals” brains and cause nervous tissue inflammation.(30). Oxidative stress, which leads to lipid and protein peroxidation, neuroinflammation, and damage to DNA and RNA, could be a contributing factor to neuronal damage and the development of autism. This hypothesis is reinforced by the current study’s findings that autistic patients exhibit an elevated oxidative stress index (OSI).

Additionally, we utilized receiver operating characteristic (ROC) to study the performance of DNA damage and DNA repair. The analysis showed a highly variable diagnostic performance for ASD. Among the biomarkers, DNA repair (OGG1) stood out as the most promising, with a high AUC indicating strong discriminatory power in differentiating ASD from non-ASD cases. While 8-oxoGuo also had a moderate predictive value, its sensitivity or specificity limitations reduce its clinical reliability when used alone.

The results suggest that OGG1 shows great potential for being included in an ASD biomarker panel, whereas 8-oxoGuo might have limited diagnostic utility. To improve sensitivity and specificity, a combined biomarker approach could be necessary, overcoming the limitations of individual markers and enhancing the accuracy of ASD diagnosis. Therefore, incorporating OGG1 as a central element in a multi-biomarker panel is advised for future ASD diagnostic methods.

However, the main limitation of our study is the small sample size, making it challenging to draw definitive conclusions about the diagnostic utility of 8-OHdG and OGG1 in autistic children. Only a limited number of biochemical analytes were evaluated. Future studies should include a broader range of analytes to provide a more comprehensive understanding of oxidative DNA damage and repair mechanisms in ASD. Additionally, ethical considerations limited the types of data we could collect, potentially impacting the scope of our findings.

## Conclusion

Our study indicates that children with autism show higher levels of intrinsic oxidative DNA damage compared to non-autistic children. The increased 8-OHdG levels in these children, along with their correlation to the severity of ASD, and the potential benefits of OGG1, bring up significant questions about the role of oxidative DNA damage in the development and progression of ASD. Additionally, the results from the ROC analysis suggest that 8-OHdG and OGG1 could be useful biomarkers for diagnosing ASD.

## Data Availability

All data produced in the present study are available upon reasonable request to the authors

https://su.edu.krd/.com

## Author contributions

The authors confirm responsibility for the following: conceptualizing and designing the study, collecting and analyzing the data, interpreting the results, and preparing the manuscript.

## Data availability statement

The data generated and examined in this study can be obtained by contacting the corresponding author upon request.

## References

1. Hirota T, King BH. Autism spectrum disorder: a review. Jama. 2023;329(2):157–68.

2. Zeidan J, Fombonne E, Scorah J, Ibrahim A, Durkin MS, Saxena S, et al. Global prevalence of autism: A systematic review update. Autism research. 2022;15(5):778–90.

3. Fombonne E, MacFarlane H, Salem AC. Epidemiological surveys of ASD: advances and remaining challenges. Journal of autism and developmental disorders. 2021;51:4271–90.

4. Steullet P, Cabungcal J, Coyle J, Didriksen M, Gill K, Grace A, et al. Oxidative stress-driven parvalbumin interneuron impairment as a common mechanism in models of schizophrenia. Molecular psychiatry. 2017;22(7):936–43.

5. Zaky EA, Abd Elhameed SA, Ismail SM, Eldamer NM, Abdelaziz AW. Analysis of urinary 8-hydroxy-2-deoxyguanosine as a biomarker of oxidative DNA damage in pediatric children with autism spectrum disorder. Research in Autism Spectrum Disorders. 2023;102:102129.

6. Whitaker AM, Schaich MA, Smith MS, Flynn TS, Freudenthal BD. Base excision repair of oxidative DNA damage: from mechanism to disease. Frontiers in bioscience (Landmark edition). 2017;22:1493.

7. Graille M, Wild P, Sauvain J-J, Hemmendinger M, Guseva Canu I, Hopf NB. Urinary 8-OHdG as a biomarker for oxidative stress: a systematic literature review and meta-analysis. International journal of molecular sciences. 2020;21(11):3743.

8. Al-Taie A, Sancar M, Izzettin FV. 8-Hydroxydeoxyguanosine: A valuable predictor of oxidative DNA damage in cancer and diabetes mellitus. Cancer: Elsevier; 2021. p. 179–87.

9. Kilicaslan F, Ayaydin H, Celik H, Kutuk MO, Kandemir H, Koyuncu I, et al. Antineuronal antibodies and 8-OHdG an indicator of cerebellar dysfunction in autism spectrum disorder: a case–control study. Psychiatry and Clinical Psychopharmacology. 2019;29(4):840–6.

10. Gohil D, Sarker AH, Roy R. Base excision repair: mechanisms and impact in biology, disease, and medicine. International Journal of Molecular Sciences. 2023;24(18):14186.

11. Giorgio M, Dellino GI, Gambino V, Pelicci PG. On the epigenetic role of guanosine oxidation. Redox Biology. 2020;29:101398.

12. Fleming AM, Zhu J, Ding Y, Esders S, Burrows CJ. Oxidative modification of guanine in a potential Z-DNA-forming sequence of a gene promoter impacts gene expression. Chemical research in toxicology. 2019;32(5):899–909.

13. Zhong Y, Zhang X, Feng R, Fan Y, Zhang Z, Zhang QW, et al. OGG1: An emerging multifunctional therapeutic target for the treatment of diseases caused by oxidative DNA damage. Medicinal Research Reviews. 2024;44(6):2825–48.

14. Bhatia S, Arslan E, Rodriguez-Hernandez LD, Bonin R, Wells PG. DNA damage and repair and epigenetic modification in the role of oxoguanine glycosylase 1 in brain development. Toxicological Sciences. 2022;187(1):93–111.

15. Tanner A, Dounavi K. The emergence of autism symptoms prior to 18 months of age: A systematic literature review. Journal of autism and developmental disorders. 2021;51(3):973–93.

16. Posar A, Visconti P. Autism in 2016: the need for answers☆. Jornal de pediatria. 2017;93:111–9.

17. Sies H, Jones DP. Reactive oxygen species (ROS) as pleiotropic physiological signalling agents. Nature reviews Molecular cell biology. 2020;21(7):363–83.

18. Hussain F, Kayani HUR. Aging-Oxidative stress, antioxidants and computational modeling. Heliyon. 2020;6(5).

19. Giese B. Long-distance electron transfer through DNA. Annual review of biochemistry. 2002;71(1):51–70.

20. Valavanidis A, Vlachogianni T, Fiotakis C. 8-hydroxy-2′-deoxyguanosine (8-OHdG): a critical biomarker of oxidative stress and carcinogenesis. Journal of environmental science and health Part C. 2009;27(2):120–39.

21. Ock C-Y, Kim E-H, Choi DJ, Lee HJ, Hahm K-B, Chung MH. 8-Hydroxydeoxyguanosine: not mere biomarker for oxidative stress, but remedy for oxidative stress-implicated gastrointestinal diseases. World journal of gastroenterology: WJG. 2012;18(4):302.

22. de Sousa MML, Ye J, Luna L, Hildrestrand G, Bjørås K, Scheffler K, et al. Impact of oxidative DNA damage and the role of DNA glycosylases in neurological dysfunction. International Journal of Molecular Sciences. 2021;22(23):12924.

23. Sidlauskaite E, Gibson JW, Megson IL, Whitfield PD, Tovmasyan A, Batinic-Haberle I, et al. Mitochondrial ROS cause motor deficits induced by synaptic inactivity: Implications for synapse pruning. Redox biology. 2018;16:344–51.

24. Bhatia S. The role of oxoguanine glycosylase 1 (OGG1) as a DNA damage repair enzyme and epigenetic modifier in mitigating the neurodevelopmental disorders initiated by physiological and ethanol-enhanced levels of reactive oxygen species: University of Toronto (Canada); 2020.

25. Porokhovnik L, Kostyuk S, Ershova E, Stukalov S, Veiko N, Korovina NY, et al. The maternal effect in infantile autism: elevated DNA damage degree in patients and their mothers. Biochemistry (Moscow) Supplement Series B: Biomedical Chemistry. 2016;10:322–6.

26. Frey BN, Andreazza AC, Kunz M, Gomes FA, Quevedo J, Salvador M, et al. Increased oxidative stress and DNA damage in bipolar disorder: a twin-case report. Progress in Neuro-Psychopharmacology and Biological Psychiatry. 2007;31(1):283–5.

27. Ming X, Stein T, Brimacombe M, Johnson W, Lambert G, Wagner G. Increased excretion of a lipid peroxidation biomarker in autism. Prostaglandins, leukotrienes and essential fatty acids. 2005;73(5):379–84.

28. James SJ, Cutler P, Melnyk S, Jernigan S, Janak L, Gaylor DW, et al. Metabolic biomarkers of increased oxidative stress and impaired methylation capacity in children with autism. The American journal of clinical nutrition. 2004;80(6):1611–7.

29. Frossi B, De Carli M, Daniel KC, Rivera J, Pucillo C. Oxidative stress stimulates IL-4 and IL-6 production in mast cells by an APE/Ref-1-dependent pathway. European journal of immunology. 2003;33(8):2168–77.

30. Theoharides TC, Kempuraj D, Redwood L. Autism: an emerging ‘neuroimmune disorder’in search of therapy. Expert Opinion on Pharmacotherapy. 2009;10(13):2127–43.

